# Diagnostic and monitoring utilities of saliva for SARS-CoV-2

**DOI:** 10.1101/2020.12.07.20244681

**Authors:** AbdulKarim AbdulRahman, Ahmed AlBastaki, Abdulla AlAwadhi, Asmaa Alwazaan, Manaf AlQahtani

**Author notes:** Corresponding author: Manaf AlQahtani.

## Abstract

**Introduction:** Nasopharyngeal (NP) swab is an invasive procedure that is difficult to perform in pediatric cases and those with special needs. On the other hand, saliva has been a proposed sample given the ease of collection, comfort and the ability to self-collect. The research project aims to study the presence of SARS-CoV-2 in the saliva of suspected COVID-19 patients in comparison to its presence in NP swabs.

**Methodology:** A cross-sectional study was conducted in October 2020 in COVID19 clinic in the Bahrain Defense Force Hospital. The study compared the presence of SARS-CoV2 by PCR in saliva samples to nasopharyngeal samples. COVID-19 Clinic tests symptomatic, staff, close contacts and pre-operation patients.

**Results:** The saliva PCR has shown a sensitivity of 72.9% (95% CI: 58.2% - 84.7%) and a specificity of 98.8% (95% CI: 97.8% - 99.4%). The PPV was 74.5% (95% CI 59.7% to 86.1%) and the NPV was 98.6% (95% CI 97.7% to 99.3%). Kappa coefficient of agreement between saliva and NP was 0.723 (95% CI 0.62 to 0.82, p<0.001). Moreover, when restricting cases to symptomatic only, the sensitivity of saliva increased to 86.7% (95% CI 59.5% to 98.3%) while specificity remained high at 97.2%.

**Conclusion:** The findings of the study suggest that saliva samples have the potential to be used as a screening tool for SARS-CoV-2, especially in symptomatic individuals. This is especially important when it is difficult to collect NP samples. Saliva samples are however at risk of producing more false negative tests.

## Introduction

The first case of novel corona virus (SARS-CoV-2) was discovered in late December 2019; a new string of the disease that causes coronavirus disease 2019 (COVID-19), which occurred in Wuhan city, China. The disease has spread rapidly across the globe that it was declared by the world health organization (WHO) as a pandemic. [1-4]. COVID-19 is an upper respiratory tract infection that is mainly spread through saliva droplets and nasal discharges while coughing or sneezing [5]. The disease symptoms include fever, cough, shortness of breath, myalgias, fatigue, loss of smell and taste [6]. Due to the high transmission rate of the disease, many countries are embracing strict activity restrictions and implemented lockdowns and curfews in an attempt to reduce the infection rate. Detection and isolation of infected individuals are essential actions in de-escalating the disease transmission [7,8].

Variety of SARS-CoV-2 detection methods are available;, nasopharyngeal swab (NP) has been the preferred specimen for polymerase chain reaction (PCR) testing due to its high detection accuracy [2,9,10]. The NP swab has several disadvantages; its invasive, painful, aerosolizing and increases risk of exposure by the healthcare worker. On the other hand, saliva has been a proposed as a diagnostic sample given the ease of collection, comfort and the ability to self-collect. The possibility of self-collection reduces the risk of exposure of healthcare workers and hence can be a safer option [11]. It was proposed that there are at least three pathways for SARS-CoV-2 to be present in the saliva. First, the virus enters the oral cavity from the upper and lower respiratory tract through the liquid droplets that are frequently exchanged between these tracts. Secondly, SARS-CoV-2 in the blood can access the oral cavity via crevicular fluid, an oral cavity-specific exudate that contains local proteins derived from extracellular matrix and serum-derived proteins Third, is by salivary gland infection, with subsequent release of particles in saliva via salivary ducts. It is essential to point out that salivary gland epithelial cells can be infected by SARS-CoV a short time after infection in rhesus macaques, suggesting that salivary gland cells could be a pivotal source of this virus in saliva [12]. However, there have been conflicting studies on the presence of SARS-CoV2 virus in the saliva sample of infected patients [1-4,8,13]. Thus, this research project aims to study the presence of SARS-CoV-2 in the saliva of suspected COVID-19 patients in comparison to its presence in NP swabs. This can help guide testing saliva as a screening tool for COVID-19.

### Objectives

Determining the presence of SARS-CoV-2 by RT-PCR in the saliva compared to its presence in NP swabs. This can assess the performance of using a saliva sample as a screening tool for detecting SARS-CoV-2

## Methods

Study Design. A cross-sectional study comparing the presence of SARS-CoV2 in NP swab and saliva samples in cases presenting to COVID-19 Clinic in the Bahrain defense force hospital. COVID-19 Clinic tests symptomatic individuals, staff, close contacts and pre-operation patients.

### Case Diagnosis

All cases diagnosed as positive for SARS-CoV-2 were based on RT-PCR tests of nasopharyngeal samples. The nasopharyngeal samples were transferred to a viral transport media immediately after collection and transported to a COVID-19 laboratory for testing. The RT-PCR tests were conducted using Thermo Fisher Scientific (Waltham, MA) Invitrogen on the Applied Biosystems (Foster City, CA) 7500 Fast Dx RealTime PCR Instrument. The assay targeted the E gene. If positive, the sample was confirmed by RdRP and N genes. The E gene CT value was reported and used in this study. CT Values >40 were considered negative.

### Saliva sample collection

- Patient were instructed not to drink, smoke or chew gum immediately beforehand.
- at least 2.5ml of saliva sample was collected in saliva collection tube (not measuring froth.)
- The patient will be instructed to spit into the tube.
- Saliva production can be helped by placing the tip of the tongue behind the front teeth.
- Saliva sample can be kept in fridge up to 72hours.

### Saliva sample processing

Saliva samples were testing using PCR. The PCR protocol followed the same protocol used for the Nasopharyngeal sample

1. If the saliva sample was mobile in nature:
  a. Take directly 200 microlitre from sample tube to RNA extraction plate and the proceed to PCR as per protocol
2. If the saliva sample is viscous in nature:
  a. Take 200 microliter sample from sample tube with Pasteur pipette to an Eppendorf tube
  b. Add 800 microliter of viral transport medium
  c. Vortex vigorously and centrifuge for 5 minutes at 2000rpm.
  d. Transfer 200 microlitre of supernatant into the extraction plate for RNA extraction and then proceed to PCR as per protocol

### Data collection and sample size

A total of 1009 patients’ samples were analyzed. Patients involved in the study had their own identifier, NP and Saliva collection date, result, and Ct values. The result was considered positive if three gene targets (E gene, ORF1ab gene and RDRP gene) were detected by RT-PCR. The cycle threshold (Ct) values >40 was considered negative. Samples positive for one or two targets were considered equivocal.

### Statistical analysis

Sensitivity, specificity, positive and negative predictive values and their 95 % confidence intervals (CI) were calculated to assess diagnostic performance. Agreement between the NP sample and saliva sample for the virus detection ability was assessed using Cohen’s Kappa (κ coefficient). Spearman correlation was used to assess the correlation between the cycle threshold (Ct) values of both the Saliva and NPS PCRs. Bland-Altman analysis was used to compare the Ct values between NPS and saliva. Differences in continuous variables were compared using a paired two sample t-test. All P-values were two-sided and P < 0.05 was considered significant. Statistical analyses were carried out using STATA 15.1

### Ethical Approval

The protocol and manuscript for this study were reviewed and approved by the Research and Research Ethics Committee in the Bahrain Defense Force Hospital (Approval Code: BDF/R&REC/2020-494). All methods and retrospective analysis of data was approved by the Committee, and carried out in accordance with local and international guidelines and regulations. Informed consent was waived by the Research and Research Ethics Committee.

## Results

A total of 1019 participants’ samples were collected. Two samples were collected from each participant, NP and saliva samples.

The Nasopharyngeal PCR resulted in 48 (4.7%) positive results, 962 (94.4%) negative and 9 equivocal (0.9%). The Saliva PCR was positive for 47 (4.6%) cases, negative for 962 (94,4) and missing for 10 (1%) cases. Cases with missing or equivocal results were excluded. 1009 sample pairs were included in the analysis

The prevalence of SARS-CoV-2 infection by NP PCR within the studied cases was 4.7% (48/1009). Table 1 shows a 2×2 contingency table of the saliva and NP PCR results.

**Table 1:** 2×2 contingency table of the saliva and NP PCR results.

|  | NP PCR+ | NP PCR- | Total |
| --- | --- | --- | --- |
| Saliva PCR+ | 35<br>True positive | 12<br>False positive | 47 |
| Saliva PCR- | 13<br>False negative | 949<br>True negative | 962 |
| Total | 48 | 961 | 1009 |

The saliva PCR has shown a sensitivity of 72.9% (95% CI: 58.2% −84.7%) and a specificity of 98.8% (95% CI: 97.8% −99.4%). The positive predictive value was 74.5% (95% CI 59.7% to 86.1%) and the negative predictive value was 98.6% (95% CI 97.7% to 99.3%). Kappa coefficient of agreement was 0.723 (95% CI 0.62 to 0.82, p<0.001) The diagnostic assessment is shown in table 2.

**Table 2:** Diagnostic assessment of saliva sample by PCR.

| Variable | Value | 95% Confidence Interval |  |
| --- | --- | --- | --- |
| Prevalence | 4.8% | 3.5% | 6.26% |
| Sensitivity | 72.9% | 58.2% | 84.7% |
| Specificity | 98.8% | 97.8% | 99.4% |
| Likelihood Ratio (+) | 58.4 | 32.4 | 105 |
| Likelihood Ratio (-) | 0.274 | 0.172 | 0.436 |
| Positive Predictive Value | 74.5% | 59.7% | 86.1% |
| Negative Predictive Value | 98.6% | 97.7% | 99.3% |
| Kappa coefficient | 72.4% | 62% | 82.7% |

To adjust for symptom status, a restricted analysis was conducted on symptomatic cases only. The sensitivity of saliva increased to 86.7% (95% CI 59.5% to 98.3%). Specificity remained high at 97.2%. Table 3 shows the 2×2 contingency table in the symptomatic sample, other assessment indicators are seen in table 4.

**Table 3:** 2×2 contingency table of the saliva and NP PCR results in symptomatic patients.

|  | NP PCR+ | NP PCR- | Total |
| --- | --- | --- | --- |
| Saliva PCR+ | 13<br>True positive | 2<br>False positive | 15 |
| Saliva PCR- | 2<br>False negative | 70<br>True negative | 72 |
| Total | 15 | 72 | 87 |

**Table 4:** Diagnostic assessment of saliva sample by PCR in symptomatic patients.

| Variable | Value | 95% Confidence Interval |  |
| --- | --- | --- | --- |
| Prevalence | 17% | 10% | 26.8% |
| Sensitivity | 86.7% | 59.5% | 98.3% |
| Specificity | 97.2% | 90.3% | 99.7% |
| Likelihood Ratio (+) | 31.2 | 7.84 | 124 |
| Likelihood Ratio (-) | 0.137 | 0.0377 | 0.499 |
| Positive Predictive Value | 86.7% | 59.5% | 98.3% |
| Negative Predictive Value | 97.2% | 90.3% | 99.7% |
| Kappa coefficient | 83.9% | 68.6% | 99.2% |

The NP samples had a median Ct value of 27.5, while the saliva had a Ct value of 28.8. Figure 1 shows a boxplot comparing Ct values from saliva and NP samples.

**Figure 1:**
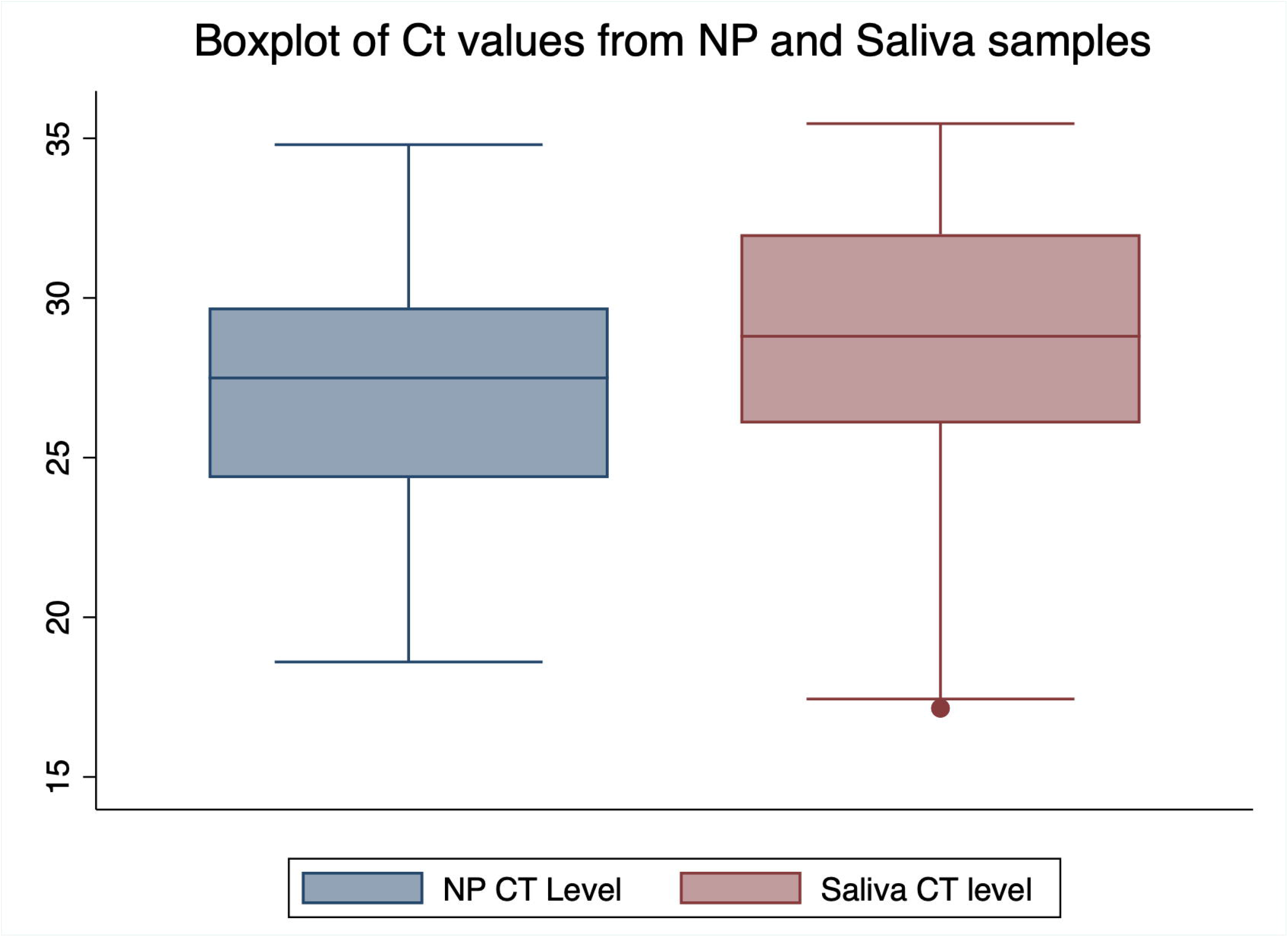
boxplot comparing Ct values from saliva and NP samples.

A paired t-test was used to compare saliva Ct value to NP Ct value in cases diagnosed using both samples (n= 35). The mean value of the saliva Ct level was 26.897 compared to 26.961 for NP CT level with a mean difference of 0.06; the difference was nonsignificant (p=0.9)

Bland-Altman analysis performed on Ct values obtained from NP and saliva samples demonstrated similar close agreement (figure 2). The Ct values from saliva and NP samples were positively correlated using spearman coefficient, however this correlation was non-significant (r= 0.3 p= 0.08). Figure 3 shows the Ct value correlation between saliva and NP samples.

**Figure 2:**
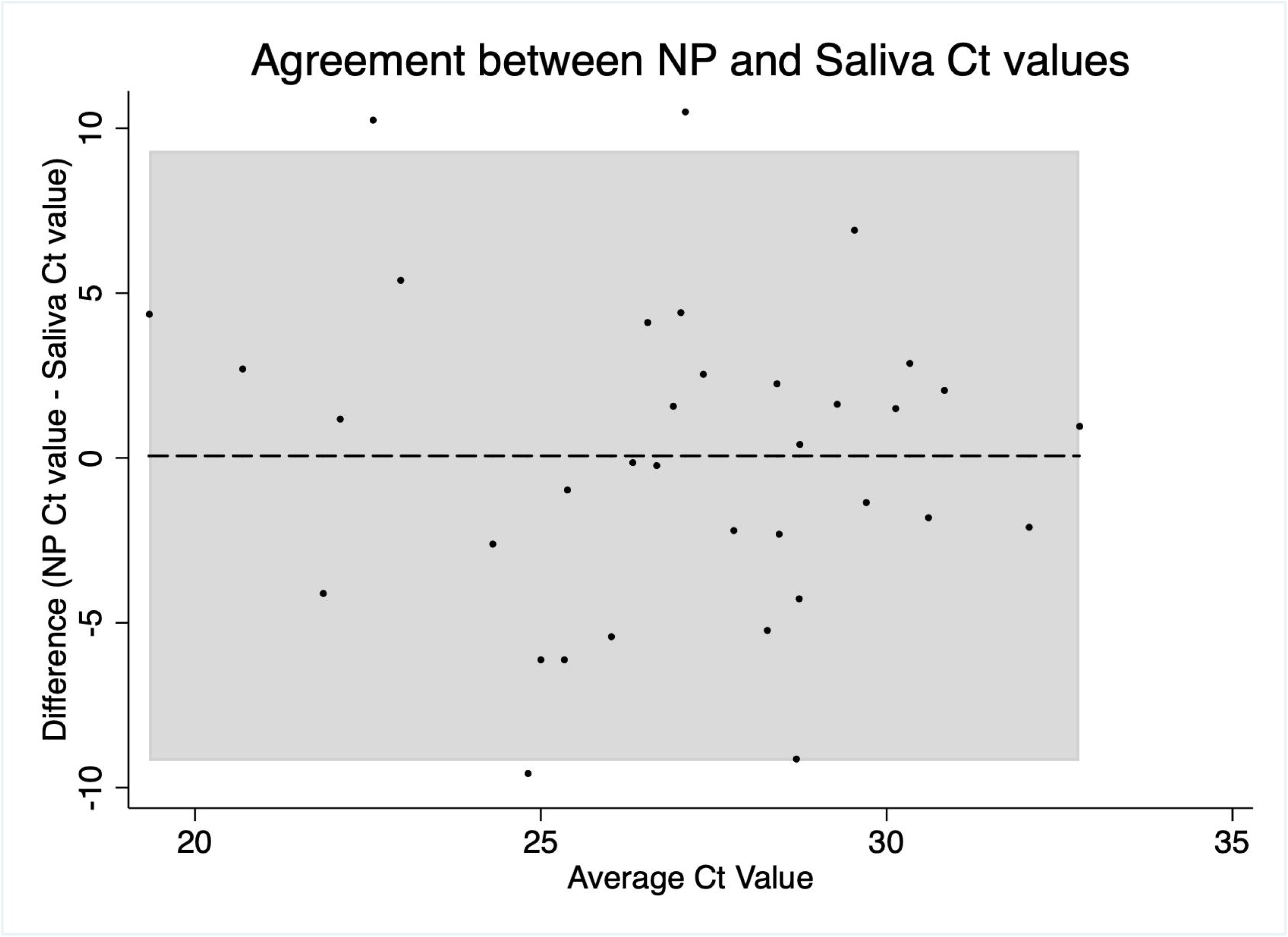
Bland-Altman analysis performed on Ct values obtained from NP and saliva samples.

**Figure 3:**
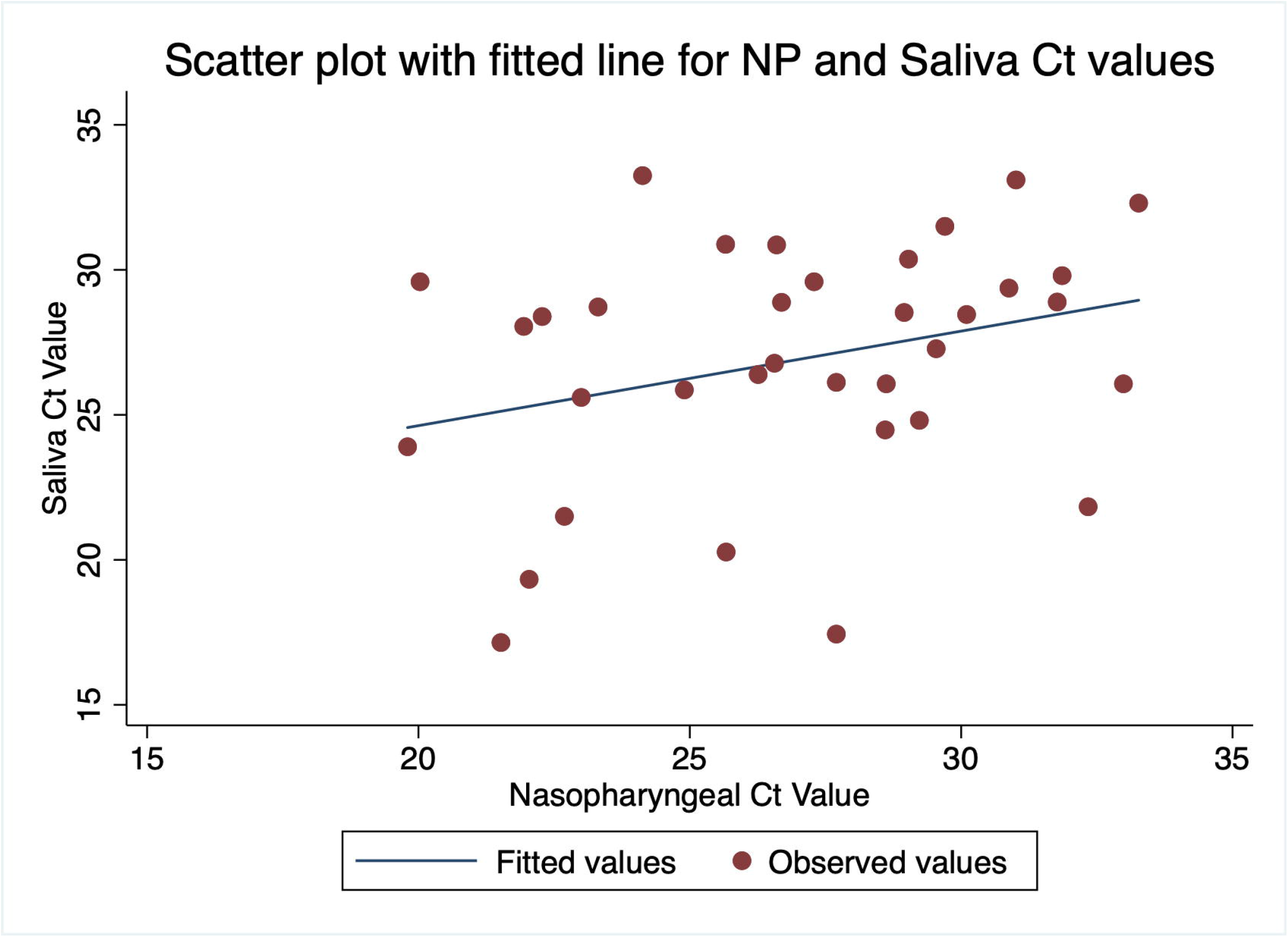
Ct value correlation between saliva and NP samples.

## Discussion

The NP swab is considered the most reliable for PCR testing and diagnosis of SARS-CoV-2 infection [14]. NP is an invasive procedure, it is reported that this technique is uncomfortable, and it induces coughing, sneezing, and causes aerosolization which increases the risk for infection transmission. The collection of NP samples can also cause nasal bleeding [1]. It is even more difficult to perform NP swab in pediatric cases and those with special needs. This difficulty can lead to improper sampling and inaccurate diagnosis. Furthermore, patient can be reluctant or hesitant to undergo testing by NP sample given its invasiveness. Saliva testing is noninvasive, comfortable and can be performed by non-medical individuals with no intra or post-procedure complications. It could also be self-collected, without risk of close contact. Several studies examined the accuracy of saliva in detecting SARS-CoV-2. The findings varied in terms of agreement.

Wyllie et al. demonstrated that saliva detected 81% of symptomatic admitted cases with COVID19 [15]. A crosse-sectional study conducted in Kuwait by Altawalah et al showed that saliva samples tested by PCR had a sensitivity of 83.43% in diagnosing symptomatic COVID19 cases in comparison to NP samples [14]. Another study by Williams et al. also reported similar findings [16]. The sensitivity of saliva sample in our study was 72.9%. This was slightly less when compared to other studies. This could be explained by the studied sample of cases. The cases within our study included a heterogenous sample of symptomatic, asymptomatic, close contacts, staff screening and pre-operative screening. The presence of asymptomatic positive individuals within our sample can explain the differences in the reported findings. As these individuals might have low viral loads. [17]

The sensitivity of the saliva sample increased to 86.7% when restricted to symptomatic cases only. This is in agreement with the majority of the reports reference above.

Yokota et al reported 86% sensitivity in using saliva samples to detect SARS-CoV-2 in asymptomatic individuals. However, the study included a full cohort of asymptomatic close contact to confirmed cases, who are considered a high-risk category [18].

The variation in the diagnostic performance of saliva samples can be due to different collection methods. Chen et al demonstrated that only 30% of infected cases were detected using saliva [19]. Chen et al reported the salivary detection rate based on pure saliva fluid secreted from the opening of salivary gland canals [19]. In Other studies patients were asked to cough out saliva from their throat into sterile containers, and hence the saliva samples were mainly sputum from the lower respiratory tract. The variation in collection technique can account to the different findings [4].

The specificity of saliva PCR was high in our studied sample, 98.8%. This can be especially important when using saliva in low-risk individuals. The specificity is in agreement with multiple manuscripts [14,16,18]. However, given the potentially lower sensitivity in asymptomatic individuals. Saliva samples can produce more false negative tests in asymptomatic cases (e.g.: pre-operative screening). It is arguable though that these cases would have lower viral load and hence could be noninfectious [20].

The findings in our study showed that saliva sample have an acceptable agreement with NP samples. The CT values between both were not different, indicating that the viral loads were similar between saliva and nasopharyngeal samples. This is in agreement with a review conducted by Fakheran et al. as they reported that there is no statistically significant difference between nasopharyngeal and saliva samples regarding viral load [4]. Similarly, Yokota et al and altawalah also had similar findings [14,18]. The correlation between saliva and Np Ct values was positive however nonsignificant in our study. This could be due to the small sample of positive cases (n=35). This could have limited the strength to detect a significant correlation. However, Procop et al reported a similar finding as they noted weak insignificant correlation between saliva and NP samples CT values. The reason for this could be due to different concentration of viral loads between patients. Patients with upper respiratory tract symptoms (e.g.: rhinorrhea) could have higher viral loads in nasopharyngeal samples, while those with oropharyngeal symptoms (e.g.: loss of taste, sore throat) could have higher viral concentrations in the salivary samples.

In our study, the positive cases in both saliva and NP samples were too small to conduct a stratified correlation analysis. Further analysis to correlate saliva and NP sample Ct values, stratified on symptom status and type of symptoms, are required to further understand this lack of correlation [20].

The study has several strengths. It included a heterogenous sample of 1009 patients. All cases had saliva and NP sample collected at the same time and analyzed in the same lab using the same standard technique. The limitations of the study included the absence of the clinical details and the low prevalence of the infection within the studied sample. The low number of positive cases limiting the strength of the study in assessing the diagnostic ability of saliva samples. Saliva samples could be best utilized for children; however, our study was performed in adult. Yet, the difference in age group is not expected to have significantly alter the results.

## Conclusion

The findings in our study adds to the rising evidence that saliva can be a reliable sample to screen for SARS-CoV-2 infection. This is especially important when it is difficult to collect NP samples. Saliva samples can be even more accurate when used in symptomatic individuals. Saliva samples are however at risk of producing more false negative tests.

## Data Availability

Available on reasonable request to corresponding author

## Declarations

### Ethics approval and consent to participate

The study was approved by the National COVID-19 Research and Ethics Committee.

### Consent for publication

All authors gave their consent for publication.

### Availability of data and materials

All the data for this study will be made available upon reasonable request to the corresponding author.

### Author contributions

AW gathered the data and supervised the laboratory team. AA and AIA analysed the data. AA, AB, AIA wrote the manuscript. AA and MQ interpreted data and edited the manuscript. All authors reviewed and approved the final version of the manuscript. Manaf Alqahtani is the guarantor of this work.

## Acknowledgment

We would like to thank our colleagues from the Bahrain defense force hospital for their cooperation to perform this research project. We would like to send our appreciation to the clinic staff and the laboratory staff in the hospital.

